# Patient-reported diagnostic pathways and disclosure experiences in autosomal dominant polycystic kidney disease (ADPKD)

**DOI:** 10.64898/2026.02.06.26345735

**Authors:** Malik Djadda, Fadi Haidar, Patrick Guirchoun, Sandra Sarthou-Lawton, Sylvie Coscoy, Dominique Joly

## Abstract

**Background and objectives:** Diagnostic disclosure practices for autosomal dominant polycystic kidney disease (ADPKD) vary and may influence patient experience and linkage to nephrology care. We characterized patient-reported diagnostic pathways, perceived timing, and disclosure experiences in the PK-DIAG survey.

**Design, setting, participants, and measurements:** Cross-sectional web-based survey in France (February 2019–August 2024) among adults with self-reported ADPKD, disseminated via patient organizations. Primary outcomes were poor tact (tact score 0–1 on a 0–5 scale) and very negative diagnostic disclosure experience (overall score 0–1 on a 0–5 scale). Multivariable logistic regression used complete-case analyses.

**Results:** Among 1,021 respondents, diagnosis was commonly disclosed outside nephrology care; 49% reported disclosure by a radiologist. Poor tact was reported by 25% and was associated with radiologist (vs nephrologist) disclosure (adjusted odds ratio 2.50; 95% confidence interval 1.57–3.98). Very negative experience was reported by 29% and was associated with poor tact (adjusted odds ratio 4.55; 95% confidence interval 2.90–7.12) and information perceived as insufficient/unclear/inaccurate (adjusted odds ratio 2.52; 95% confidence interval 1.53–4.15). Most participants perceived diagnosis as timely (67%), while 18% perceived it as too early and 15% as too late. Distress (36%) or unmet psychological needs (40%) in the immediate post-disclosure period was common.

**Conclusions:** ADPKD diagnostic disclosure often occurred outside dedicated nephrology consultations and was frequently associated with poor tact and inadequate information. These findings support structured, guideline-aligned disclosure pathways incorporating timely counseling, psychosocial support, and rapid linkage to nephrology care.

**Key Points:**

- In PK-DIAG, initial disclosure of ADPKD frequently occurred outside nephrology care, most often by radiologists in radiology settings.
- About one quarter of respondents reported poor tact and 29% reported a very negative overall diagnostic disclosure experience.
- Poor tact and information perceived as insufficient/unclear/inaccurate were strongly associated with a very negative diagnostic experience.

## Introduction

Autosomal dominant polycystic kidney disease (ADPKD) is the most common inherited kidney disease, characterized by progressive cyst development leading to kidney failure (1). Beyond the somatic burden of pain, organ enlargement, and extrarenal manifestations, ADPKD carries distinct psychosocial weight. Its hereditary nature imposes unique anxieties regarding disease transmission, family planning, and identity (2,3).

For many at-risk individuals, ADPKD remains an observed disease in relatives until a personal diagnosis marks a salient transition into the disease. As established in oncology, the quality of diagnostic disclosure is a key component of the patient experience. Better-managed disclosure has been associated with greater trust and shared decision-making, whereas poorer disclosure has been linked to greater anxiety and feelings of guilt (4).

Diagnosis typically occurs via family screening, symptom evaluation, or increasingly, as an incidental finding. The optimal timing of diagnosis remains ethically complex: while early identification allows for monitoring and lifestyle interventions, it may impose the psychosocial burden of a chronic label without immediate curative options—a “sword of Damocles” effect described in qualitative studies (5). This tension is intensifying with advances in imaging and prenatal testing.

The 2025 KDIGO guideline emphasizes that diagnosis should be accompanied by counseling on prognosis and genetics (6). However, the real-world implementation of these recommendations depends on how, where, and by whom the diagnosis is communicated. Evidence describing routine diagnostic pathways and the patient experience of disclosure remains limited.

We conducted the PK-DIAG survey to capture the diagnostic pathway from the patient perspective. We hypothesized that modifiable features of disclosure are associated with adverse patient experiences and subsequent engagement with care. Accordingly, the main objectives of this study were to (i) describe diagnostic pathways and disclosure practices, (ii) characterize the patient experience of diagnosis, and (iii) identify factors associated with poor disclosure and downstream engagement with nephrology care.

## Methods

Study design and population. PK-DIAG was a cross-sectional, web-based survey conducted in France (February 2019–August 2024) among adults (≥18 years) with self-reported ADPKD across CKD stages (including dialysis and transplant), disseminated via national patient organizations.

Questionnaire development. A 72-item questionnaire was developed from literature review and expert consensus, refined through pilot testing, reviewed by patient associations (PKD France, AIRG), and digitally pre-tested for usability (n=41).

Measures. Domains included diagnostic circumstances, disclosure experience, perceived information quality at disclosure, health status, and quality of life (ADPKD-IS; higher scores indicate greater impact).

Disclosure experience and information. Because no ADPKD-specific validated instrument exists for routine disclosure quality, tact and overall diagnostic experience were each assessed with a single-item numeric rating scale (0=worst possible, 5=best possible); exact item wording and anchor labels are provided in Supplementary Table 6. Perceived information was categorized as sufficient vs insufficient, clear vs unclear, and accurate vs inaccurate; “poor information” denotes any negative rating.

Outcomes. Primary outcomes were poor tact at disclosure (tact score 0–1 on a 0–5 scale) and a very negative diagnostic disclosure experience (global experience score 0–1 on a 0–5 scale). For regression analyses, each score was dichotomized a priori as 0–1 versus 2–5 to capture clearly negative experiences (lowest two response categories) and to facilitate interpretation of odds ratios. To document the full range of responses and potential floor/ceiling effects, we additionally report the complete 0–5 score distributions (including median [IQR] and the proportions at scores 0 and 5) in the Supplementary Appendix.

Data collection and ethics. Participation was voluntary and anonymous; no directly identifiable data were collected (GDPR-compliant). The study was approved by the AP-HP Centre IRB (CERAPHP.5, IRB #00011928).

Deduplication. Duplicate/triplicate entries were removed using a prespecified algorithm (pseudonym-based grouping and internal consistency checks) followed by manual review.

Statistical analysis. We used descriptive statistics and multivariable logistic regression with a priori covariates; each model used complete cases and reports the analytical n. To further characterize the distributional properties of the disclosure experience scores, we additionally described the full distribution of the tact score and the global diagnostic experience score across all response categories (0–5), reporting median [interquartile range], percentages by response option, and floor and ceiling effects. These descriptive analyses were performed to assess scale utilization and support the interpretability of the predefined dichotomization thresholds; detailed results are provided in Supplementary Table 7. Variables measured at the time of the survey (e.g., current mood, ADPKD-IS) were treated as exploratory adjustments and not interpreted as temporally antecedent. Analyses were performed in R (v4.3.1).

## Results

### Participant Characteristics

After removing 32 duplicate or triplicate entries, 1,021 unique responses were included in the final analysis (Table 1). Mean age at the time of the survey was 51.2 ± 14.0 years (N=797), and 531/797 (67%) were women. Among respondents with education data (N=895), 742/895 (83%) reported at least a bachelor’s degree. Kidney replacement therapy (KRT) status was available for 837 respondents; 232/837 (28%) had initiated KRT, including 60/837 (7%) receiving dialysis and 172/837 (21%) living with a functioning kidney transplant. A family history of ADPKD was reported by 639/811 (79%); among respondents with a family history who answered the index-case item (N=634), 88/634 (14%) identified themselves as the first diagnosed individual in their family. Item-level missingness was present; therefore, denominators vary across variables and are reported throughout the text and tables. Geographically, respondents were distributed across metropolitan France; the density of participation by department is illustrated in Supplementary Figure 1.

**Table 1.**
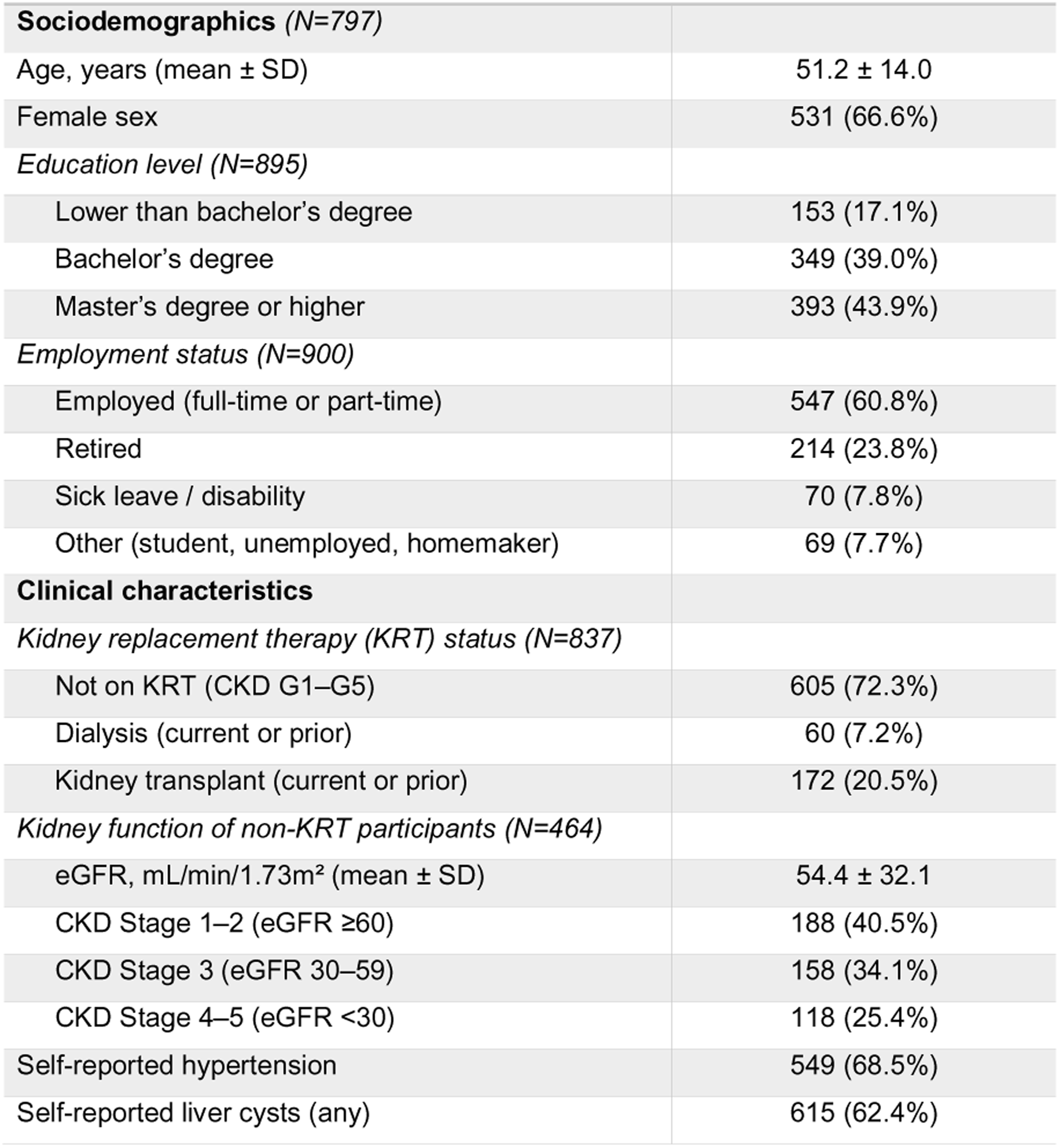
Participant characteristics. Participant characteristics (n=1,021). Values are mean ± SD or n (%), with percentages calculated among respondents with available data for each variable. Denominators vary because of item-level missingness and conditional questions. KRT, kidney replacement therapy; CKD, chronic kidney disease; eGFR, estimated glomerular filtration rate.

|  |  |
| --- | --- |
| **Sociodemographics (N=797)** |  |
| Age, years (mean ± SD) | 51.2 ± 14.0 |
| Female sex | 531 (66.6%) |
| *Education level (N=895)* |  |
| Lower than bachelor's degree | 153 (17.1%) |
| Bachelor's degree | 349 (39.0%) |
| Master's degree or higher | 393 (43.9%) |
| *Employment status (N=900)* |  |
| Employed (full-time or part-time) | 547 (60.8%) |
| Retired | 214 (23.8%) |
| Sick leave / disability | 70 (7.8%) |
| Other (student, unemployed, homemaker) | 69 (7.7%) |
| **Clinical characteristics** |  |
| *Kidney replacement therapy (KRT) status (N=837)* |  |
| Not on KRT (CKD G1–G5) | 605 (72.3%) |
| Dialysis (current or prior) | 60 (7.2%) |
| Kidney transplant (current or prior) | 172 (20.5%) |
| *Kidney function of non-KRT participants (N=464)* |  |
| eGFR, mL/min/1.73m<sup>2</sup> (mean ± SD) | 54.4 ± 32.1 |
| CKD Stage 1–2 (eGFR ≥60) | 188 (40.5%) |
| CKD Stage 3 (eGFR 30–59) | 158 (34.1%) |
| CKD Stage 4–5 (eGFR <30) | 118 (25.4%) |
| Self-reported hypertension | 549 (68.5%) |
| Self-reported liver cysts (any) | 615 (62.4%) |

### Diagnostic pathways and age at diagnosis

Diagnostic pathways are summarized in Figure 1 and Table 2. Diagnosis occurred most frequently via family screening (40.1%), followed by symptom investigation (26.6%) and incidental findings (20.7%). The median age at diagnosis was 26 [IQR 18–36] years. Diagnosis before age 18 occurred in 18.2% whereas reported prenatal or at-birth diagnosis was uncommon (1.4%).

**Figure 1.**
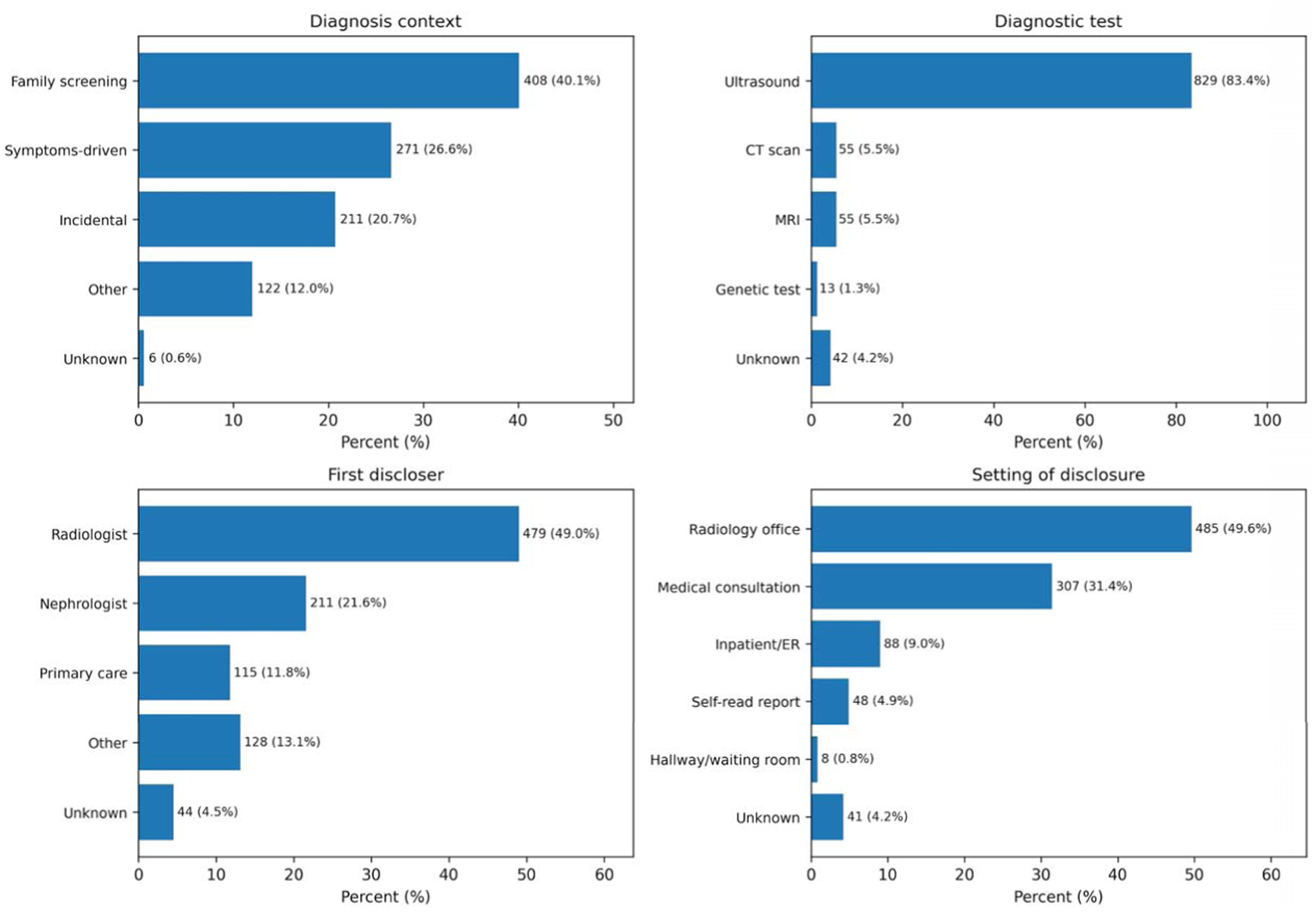
Diagnostic pathway overview. The diagrams summarize patient-reported diagnostic context (family screening, symptom-driven workup, incidental detection, other), main diagnostic test(s), first professional disclosing the diagnosis, and setting of disclosure. Values are n/N (%) based on available responses; denominators vary because of missing and conditional questions. CT, computed tomography; MRI, magnetic resonance imaging; ER, emergency room

**Table 2.** Diagnostic pathways and disclosure experience. Values are n/N (%) unless otherwise stated. Percentages are calculated on valid responses. Tact score and global experience score are single-item numeric rating scales ranging from 0 (worst possible experience) to 5 (best possible experience). Poor tact is defined a priori as a tact score 0–1; very negative experience as a global experience scores 0–1. “Poor information” is defined as information rated insufficient and/or unclear and/or inaccurate at the time of disclosure.

| Diagnostic circumstances | Value |
| --- | --- |
| **Age at diagnosis, years (mean ± SD) (N=1001)** | 28.8 ± 13.4 |
| **Early diagnosis (<18 years), (N=1001)** | 182(18%) |
| **Context of diagnosis, (N=1018)** |  |
| Family screening (asymptomatic) | 408 (40%) |
| Investigation of symptoms | 271 (27%) |
| Incidental finding | 211 (21%) |
| Other | 122 (12%) |
| Unknown | 6 (0.6%) |
| **Diagnostic modality, (N=994)** |  |
| Ultrasound | 829 (83%) |
| CT scan / MRI | 110 (11%) |
| Genetic testing | 13 (1 %) |
| Unknown | 42 (4 %) |
| **The disclosure encounter** |  |
| **First professional to disclose, (N=977)** |  |
| Radiologist | 479 (49 %) |
| Nephrologist | 211 (22%) |
| Primary care physician | 115 (12%) |
| Other specialist | 128(13%) |
| Unknown | 44 (5%) |
| **Setting of disclosure, (N=977)** |  |
| Radiology office | 485 (50%) |
| Medical consultation | 307 (31%) |
| Inpatient / Emergency room | 88 (9%) |
| Self-read report | 48 (5%) |
| Hallway / Waiting room | 8 (1%) |
| Unknown | 41(4%) |
| **Patient-Reported Experience (PREMs)** |  |
| Tact score (0–5 scale), mean ± SD (N=928) | 2.6 ± 1.6 |
| Poor Tact reported (Score 0–1), n (%) (N=928) | 229 (25%) |
| Overall experience score (0–5 scale), mean ± SD (N=927) | 2.4 ± 1.5 |
| Very negative experience (Score 0–1), n (%) (N=927) | 268 (29%) |
| Information rated as insufficient/unclear/inaccurate, (N=974) | 218 (22%) |
| Immediate psychological support: unmet need, (N=974) | 394 (40 %) |

Within the family screening cohort, women received a diagnosis earlier than men (median 20 vs. 23 years; p=0.025). The decision to screen was initiated by parents in 34.6% of cases and by respondents in 34.0%; however, parent-initiated screening was associated with a significantly earlier diagnosis (median 18 years) compared with self-referral (median 29 years). In multivariable linear regression restricted to postnatal diagnoses (age at diagnosis >0), male sex and non–family-screening diagnostic contexts were independently associated with an older age at ADPKD diagnosis (Figure 2 and supplementary Table 1). In the overall cohort (complete cases, n=770), men were diagnosed 4.22 years later than women (95% CI 2.26–6.17; p<0.001), and diagnosis during symptom-driven evaluation or incidental detection occurred approximately 6 years later than family screening (both p<0.001). A reported family history was associated with earlier diagnosis (−5.29 years; 95% CI −7.71 to −2.88; p<0.001). In the family-screening subgroup (n=324), screening initiation was strongly associated with age at diagnosis: compared with parent-initiated screening, self-initiated screening was associated with a ∼10-year older age at diagnosis (+10.23 years; 95% CI 7.35–13.11; p<0.001).

**Figure 2.**
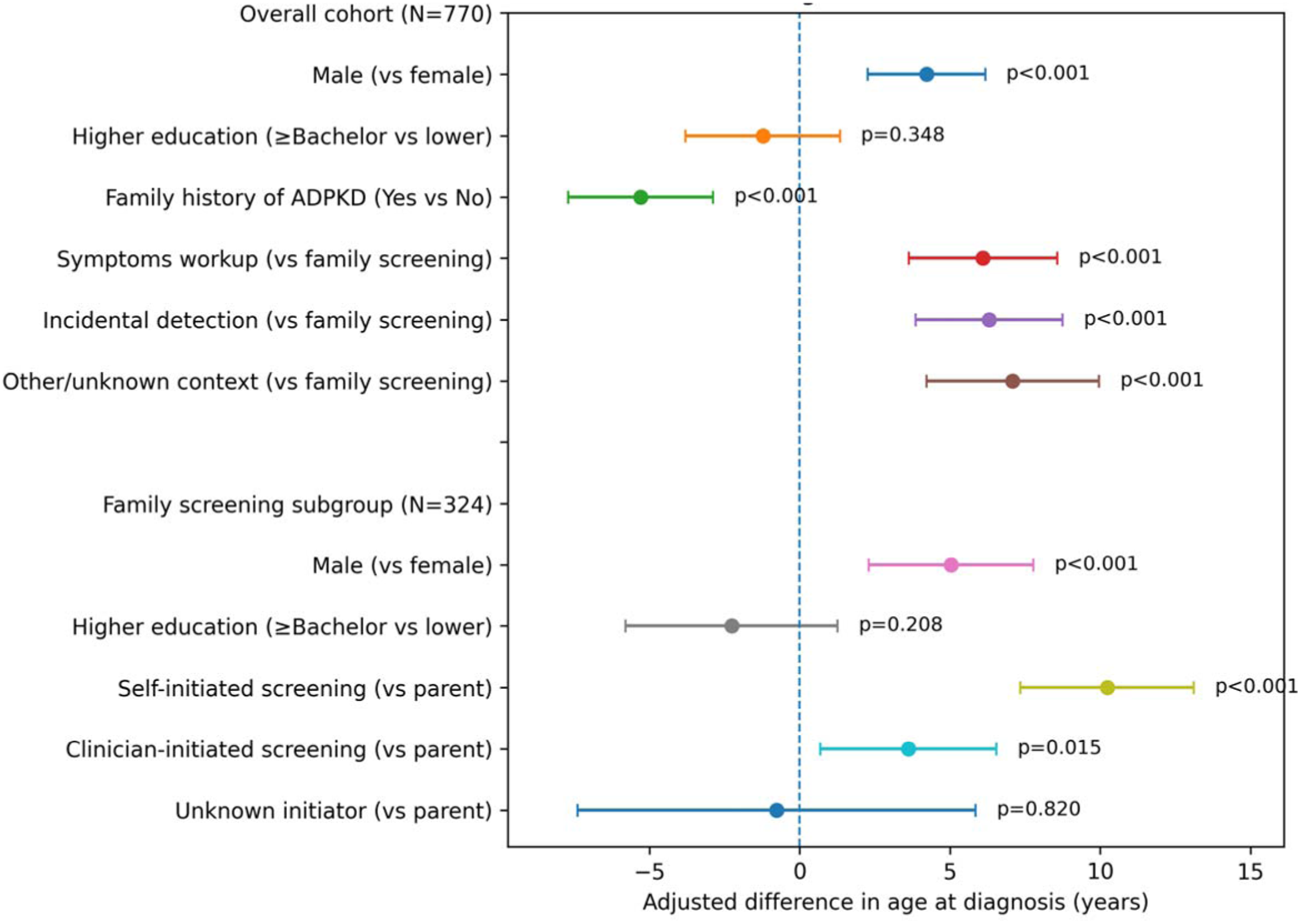
Multivariable factors associated with postnatal age at ADPKD diagnosis (linear regression) Points represent regression coefficients (β) and whiskers represent 95% confidence intervals. Coefficients reflect the adjusted mean difference in age at diagnosis (years) associated with each covariate. Upper panel: overall cohort model; lower panel: family-screening subgroup including screening initiator. Models were fitted as complete-case analyses using robust standard errors.

The disclosure experience. Ultrasound was the main diagnostic modality (83.4%). Diagnosis was disclosed by a radiologist in 49.0% of cases, often in a radiology office (49.6%). Approximately one-quarter of participants (24.5%) reported “poor tact” during disclosure (Table 2). Tact score used the full response range, with median [IQR] values of 3 [2–4] and moderate floor effects without marked ceiling effects (Supplementary Table 7). In multivariable logistic regression, poor tact (Score 0-1) was independently associated with radiologist disclosure compared with nephrologist disclosure (aOR 2.50, 95% CI 1.57–3.98), incidental diagnosis compared with family screening (aOR 2.11, 95% CI 1.36–3.27), and symptom-driven diagnosis compared with family screening (aOR 1.62, 95% CI 1.04–2.52). Age at diagnosis, sex, and educational level were not independently associated with poor tact in this model (Figure 3 and Supplementary Table 2).

**Figure 3.**
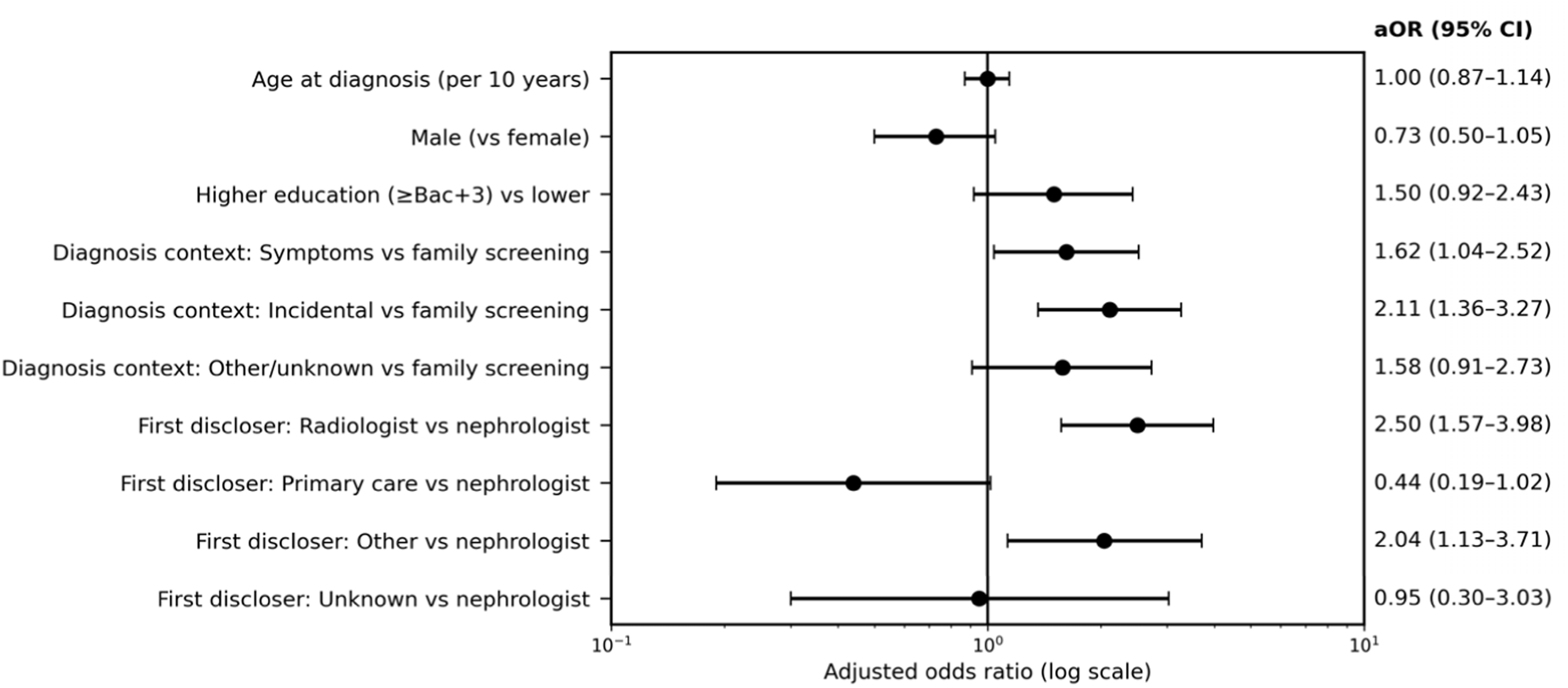
Multivariable factors associated with poor tact at ADPKD diagnosis (logistic regression) Forest plot of adjusted odds ratios (aOR) with 95% confidence intervals for poor tact (tact score 0–1 on a 0–5 scale). Points and whiskers (black) indicate aORs and 95% confidence intervals; numeric aOR (95% CI) values are displayed on the plot. Model fitted as a complete-case analysis (n=803). Reference categories are indicated in the figure.

### Emotional response: fears and feelings

The most common fears at diagnosis were kidney failure (61.3%) and transmission to children (47.5%) (Supplementary Table 3). Feelings experienced in the hours/days following diagnosis reported were uncertainty (31.3%), indifference (30.0%), anxiety/stress (27.9%), shock (20.7%), and sadness/depression (14.3%) (Supplementary Table 4). A composite distress outcome (anxiety/stress and/or sadness/depression) was present in 36.2% and was independently associated with poor tact (aOR 1.93 [1.29–2.90]); poor information (aOR 1.99 [1.30–3.04]); older age at diagnosis (aOR 1.29 per 10-year increase [1.12–1.49]); and greater worry related to family history (aOR 1.52 per point [1.28–1.81]) (Supplementary Table 4).

### Overall diagnostic experience

Overall diagnostic disclosure experience was rated on a 0–5 scale (0 being worst, 5 being best; mean 2.4±1.5; Table 2). Scores used the full scale response range, with median [IQR] values of 2 [1–3], and moderate floor or ceiling effects (Supplementary Table 7). A very negative diagnostic experience (score 0–1) was reported by 29.1%. In multivariable logistic regression (n=596 complete cases), the strongest independent correlates were encounter-level factors: poor tact (aOR 4.55 [2.90–7.12]) and poor information (aOR 2.52 [1.53–4.15]). Greater worry about family history was also independently associated with very negative experience (aOR 1.37 per level [1.09–1.71]). Because several covariates were collected retrospectively or at the time of the survey (e.g., psychological support around diagnosis and current mood/ADPKD-IS), their temporality relative to the disclosure is uncertain and reverse causality cannot be excluded; we therefore treat these as exploratory adjustment variables and do not interpret them as temporally antecedent explanatory factors (Figure 4; Supplementary Table 5).

**Figure 4.**
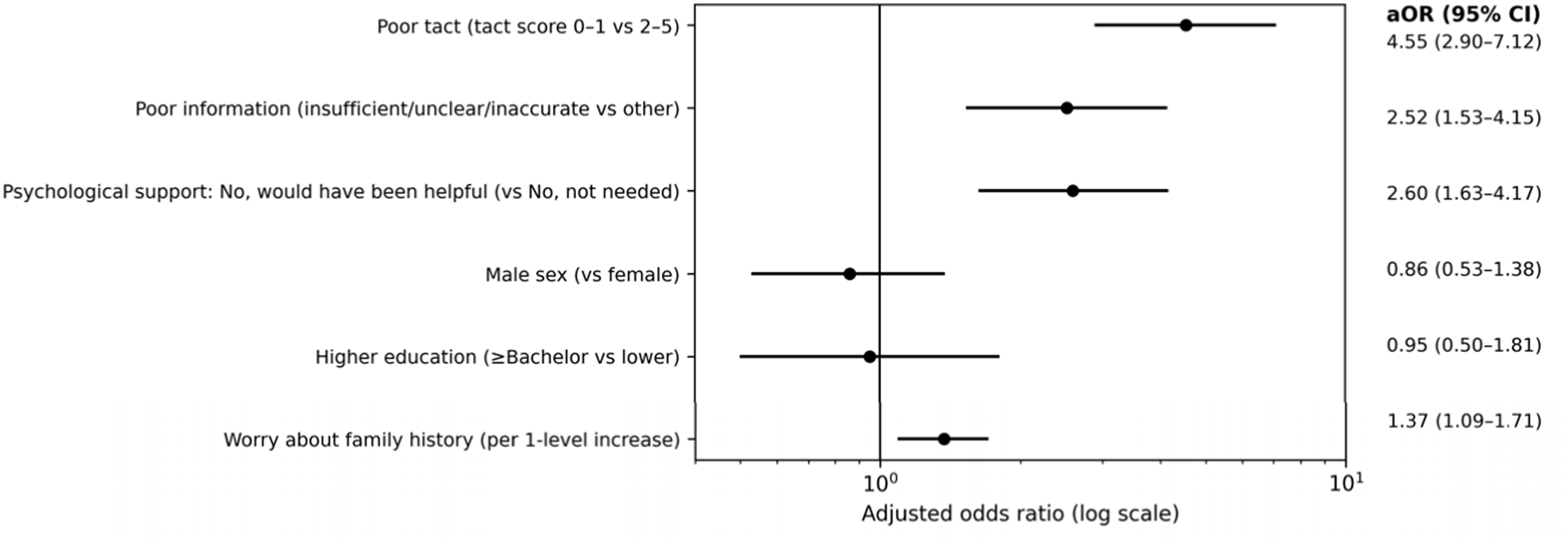
Multivariable factors associated with very negative diagnostic disclosure experience. Forest plot of adjusted odds ratios (aOR) from multivariable logistic regression for very negative experience, defined as a global diagnostic experience score 0–1 on a 0–5 scale (vs 2–5). Points and whiskers indicate aORs and 95% confidence intervals; numeric aOR (95% CI) values are displayed on the plot. Model fitted as a complete-case analysis (n=596). For clarity, this figure displays selected covariates with clear temporal anchoring at the time of disclosure; exploratory covariates measured at the time of the survey (current mood and ADPKD-IS) and the sparse category “psychological support received” are not displayed. Full model specification and estimates are provided in Supplementary Table 5.

### Perceived timing

While 67.1% felt their diagnosis occurred at the “right time,” substantial minorities felt it was “too early” (18.1%) or “too late” (14.8%). Both dissatisfied groups reported worse emotional quality of life scores (Table 3). When asked what age they considered most useful to know one is affected by ADPKD within the family, 42.7% favored 18–30 years, 33.1% favored <18 years, and 13.5% favored >30 years, while 10.6% reported no opinion. Opinions regarding the optimal age for diagnosis varied significantly by personal experience. Participants who perceived their own diagnosis as premature were more likely to favor a later age, whereas those who felt it was delayed favored an earlier age. Furthermore, the discrepancy between actual and preferred age at diagnosis was consistent with the subjective perception of timing (Table 3).

**Table 3.**
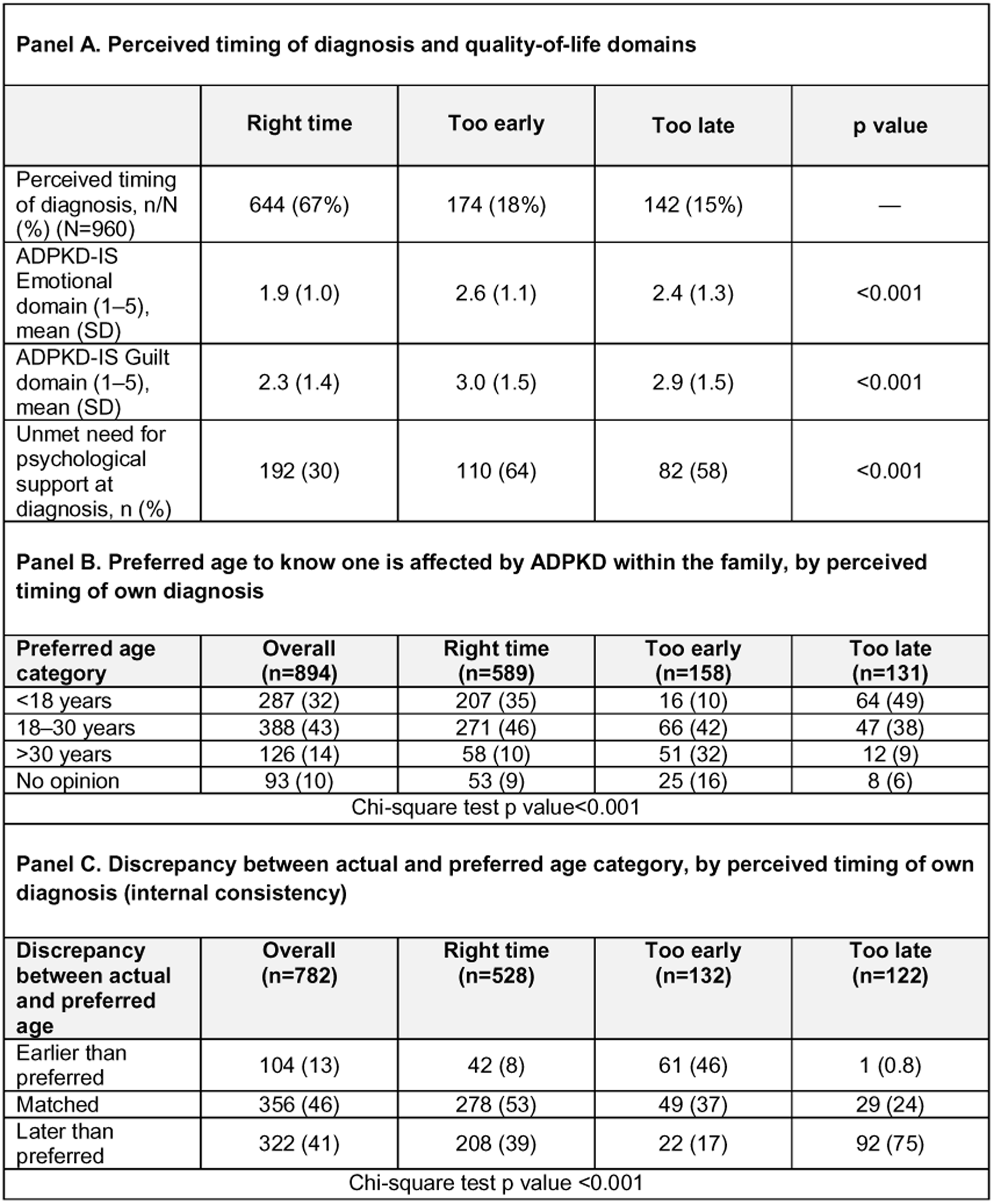
Perceived timing of ADPKD diagnosis, preferred age to know one is affected within the family, and internal consistency analyses. **(A)**Association between perceived timing of diagnosis (classified as “too early,” “right time,” or “too late”) and patient-reported outcomes. Outcomes include the proportion of patients reporting an unmet need for immediate psychological support and scores for the ADPKD-IS Emotional and Guilt domains (presented as mean ± SD). Higher ADPKD-IS scores (range 0–5) indicate greater negative impact. **(B)** Preferred age category to know one is affected by ADPKD (<18 years, 18–30 years, >30 years, or no opinion), stratified by the participant’s retrospective appraisal of their own diagnostic timing. **(C)** Internal consistency analysis comparing the subjective appraisal of timing with the calculated discrepancy between actual and preferred age of diagnosis (categorized as earlier than preferred, matched, or later than preferred). *Note: Values are n (%) unless otherwise indicated. Percentages in Panels B and C are column percentages. P values are derived from Chi-square tests*

| **Panel A. Perceived timing of diagnosis and quality-of-life domains** |  |  |  |  |
| --- | --- | --- | --- | --- |
|  | **Right time** | **Too early** | **Too late** | **p value** |
| Perceived timing of diagnosis, n/N (%) (N=960) | 644 (67%) | 174 (18%) | 142 (15%) | — |
| ADPKD-IS Emotional domain (1–5), mean (SD) | 1.9 (1.0) | 2.6 (1.1) | 2.4 (1.3) | <0.001 |
| ADPKD-IS Guilt domain (1–5), mean (SD) | 2.3 (1.4) | 3.0 (1.5) | 2.9 (1.5) | <0.001 |
| Unmet need for psychological support at diagnosis, n (%) | 192 (30) | 110 (64) | 82 (58) | <0.001 |
| **Panel B. Preferred age to know one is affected by ADPKD within the family, by perceived timing of own diagnosis** |  |  |  |  |
| **Preferred age category** | **Overall (n=894)** | **Right time (n=589)** | **Too early (n=158)** | **Too late (n=131)** |
| <18 years | 287 (32) | 207 (35) | 16 (10) | 64 (49) |
| 18–30 years | 388 (43) | 271 (46) | 66 (42) | 47 (38) |
| >30 years | 126 (14) | 58 (10) | 51 (32) | 12 (9) |
| No opinion | 93 (10) | 53 (9) | 25 (16) | 8 (6) |
| Chi-square test p value<0.001 |  |  |  |  |
| **Panel C. Discrepancy between actual and preferred age category, by perceived timing of own diagnosis (internal consistency)** |  |  |  |  |
| **Discrepancy between actual and preferred age** | **Overall (n=782)** | **Right time (n=528)** | **Too early (n=132)** | **Too late (n=122)** |
| Earlier than preferred | 104 (13) | 42 (8) | 61 (46) | 1 (0.8) |
| Matched | 356 (46) | 278 (53) | 49 (37) | 29 (24) |
| Later than preferred | 322 (41) | 208 (39) | 22 (17) | 92 (75) |
| Chi-square test p value <0.001 |  |  |  |  |

### Post-diagnosis care

After diagnostic disclosure, 38.5% of respondents reported a resolution to initiate specialist follow-up, whereas 36.9% reported no specific resolution (multi-select responses). Overall, 60 % reported seeing a nephrologist promptly after diagnosis, while 38 % reported a first nephrology consultation only years later and 2% had never or not yet consulted; thus, 40 % reported delayed or absent nephrology follow-up (Table 4 panel A).

**Table 4.** Post-disclosure linkage to nephrology care and factors associated with delayed/never nephrology follow-up. Panel A shows the timing of the first nephrology consultation after ADPKD diagnosis. Panel B displays the concordance between reported post-disclosure resolutions and the timing of nephrology follow-up. Panel C presents an exploratory multivariable logistic regression examining factors associated with delayed or absent nephrology follow-up. Notes : Delayed/never follow-up combines “years later” and “never / not yet”. Poor information defined as information rated insufficient and/or unclear and/or inaccurate at diagnosis.

| **A. Timing of first nephrology consultation after diagnosis (N=936)** |  |  |
| --- | --- | --- |
|  | **n** | **%** |
| Promptly (soon after diagnosis) | 563 | 60 |
| Delayed or never/not yet | 373 | 40 |
| Years later | 355 | 38 |
| Never / not yet | 18 | 2 |
| **B. Timing stratified by patient resolution to initiate specialist follow-up** |  |  |
| **Post disclosure resolution** | **Promptly**<br>**n (row %)** | **Delayed/never\***<br>**n (row %)** |
| Resolution to initiate specialist follow-up | 302 (84) | 59 (16) |
| No specific resolution | 129 (37) | 219 (63) |
| **C. Factors associated with delayed/never nephrology follow-up vs prompt follow-up (reference)** |  |  |
| **Predictor** | **Adjusted OR (95%**<br>**CI)** | **P value** |
| Resolution: no specific resolution | 4.3 (3.1–6.1) | <0.001 |
| Poor information quality at diagnosis | 1.84 (1.3–2.7) | 0.02 |
| Age at diagnosis (per 10-year increase) | 0.8 (0.7–0.9) | <0.001 |
| Male sex (vs female) | 1.06 (0.75–1.51) | 0.7 |
| Higher education (≥ Bachelor's degree) | 0.9 (0.6–1.4) | 0.7 |
| Diagnostic context: symptoms/driven workup (vs family screening) | 0.55 (0.5–0.8) | 0.006 |
| Diagnostic context: incidental finding/other/unknown (vs family screening) | 0.9 (0.6–1.4) | 0.7 |

Prompt nephrology follow-up was more frequently reported among participants who indicated a resolution to initiate specialist follow-up than among those who did not (83.3% vs 45.4%; Table 4 panel B), indicating a strong concordance between reported post-disclosure intentions and the reported timing of the first nephrology consultation. Given the cross-sectional design and retrospective reporting of both intentions and care trajectories, this association should be interpreted descriptively and may be affected by recall bias or reverse causality.

In exploratory multivariable analysis comparing delayed/never versus prompt nephrology follow-up, reporting no specific resolution at disclosure was strongly associated with delayed or absent follow-up (adjusted OR 4.3, 95% CI 3.1–6.1). Poor information at diagnosis was also associated with delayed/never follow-up (adjusted OR 1.8, 95% CI 1.3–2.7), whereas older age at diagnosis was associated with lower odds of delayed follow-up (adjusted OR 0.8 per 10 years, 95% CI 0.7–0.9). Compared with family screening, diagnosis occurring during a symptom-driven workup was associated with lower odds of delayed/never nephrology follow-up (adjusted OR 0.6, 95% CI 0.4–0.8), while incidental or other diagnostic contexts did not differ significantly (Table 4 panel C).

## Discussion

In this large, de-duplicated patient-reported survey, ADPKD diagnosis was often disclosed in radiology settings—frequently by radiologists—and one quarter of respondents reported poor tact. Nearly one third reported a very negative overall diagnostic disclosure experience, which was strongly associated with patient-reported disclosure tact and the perceived adequacy and clarity of information. While causal inference is not possible in this design, these findings are consistent with the hypothesis that aspects of the disclosure encounter represent actionable targets for quality improvement.

### Diagnostic pathways, family screening, and the role of family dynamics

Age at ADPKD diagnosis largely reflected pathways to care. Diagnoses made outside family screening occurred later, whereas parent-initiated screening occurred markedly earlier than self-initiated screening, consistent with cascade-screening experience in inherited conditions (7). Parent-initiated screening was associated with much earlier diagnosis, highlighting the role of family dynamics in the diagnostic pathway, a finding consistent with recent pediatric registry data from Gimpel et al. which shows stable rates of active screening in minors over time (8). Men were diagnosed later than women, likely reflecting differences in screening uptake and care-seeking rather than disease onset. These data support family-centered counseling and cascade-screening workflows to align diagnostic timing with patient preferences while minimizing psychosocial harm.

### Early and presymptomatic testing: patient ambivalence and ethical considerations

A substantial minority perceived their diagnosis as occurring at the wrong time (too early or too late), and these groups reported worse emotional and guilt-related quality-of-life domains. This is consistent with qualitative work describing substantial emotional burden even in early stages (9) and with ambivalence toward presymptomatic testing (5). Pediatric consensus statements similarly emphasize that testing in asymptomatic minors should not be routine and requires careful balancing of potential medical benefit and psychological harm (10). In PK-DIAG, fears at diagnosis were dominated by the prospect of kidney failure and transmission to offspring, emphasizing disclosure encounters that address prognosis, inheritance risk, reproductive counseling, and psychosocial support. Timing-related measures showed strong internal coherence, supporting the construct validity of these patient-reported items.

### Disclosure practices in routine care and alignment with KDIGO guidance

The 2025 KDIGO guideline emphasizes that diagnostic and predictive testing should be accompanied by counseling on benefits and potential harms, and that counseling should address implications for prognosis, inheritance, and family planning (6). Our data suggest that disclosure often occurs before such counseling is delivered, particularly when first communicated in imaging settings or after incidental findings. Radiologists play a crucial technical role but may be constrained in addressing complex genetic and prognostic issues at the time of first disclosure (4). These findings support practical, guideline-aligned strategies such as prompt referral to nephrology after imaging suspicion, standardized written information, and systematic screening for psychological distress with clear referral pathways.

### Psychosocial burden and patient priorities

The high prevalence of anxiety (27.9%) and fear of transmission (47.5%) in our cohort represents a significant burden, echoing findings on depression and psychosocial risk in ADPKD (11). Recent patient-centered research by Mustafa et al. (EMPOWER-PKD) identified “psychological effect” as a leading priority for patients, ranking it higher than mortality in terms of daily impact (4). Similarly, the SONG-PKD initiative confirmed that patients prioritize “life impacts“—such as anxiety and ability to work—well before the biological markers typically monitored by clinicians (12). Our findings validate this prioritization and show that a negative diagnostic experience is strongly linked to *modifiable* factors (tact, information). The use of ADPKD-specific patient-reported outcome measures (PROMs), such as ADPKD-IS and ADPKD-PDS, can better capture these impacts that generic tools may underestimate (13–15).

### Bridging diagnosis and specialist care

Associations between immediate post-disclosure resolutions and subsequent nephrology follow-up (Table 4) should be interpreted cautiously, as both may be related to the same underlying factors (e.g., referral pathways, disease severity at diagnosis, and family context) and reporting may be affected by retrospective appraisal. In addition, the strong alignment between reporting an intention to seek specialist care and reporting prompt nephrology follow-up may partly reflect internal consistency in self-reporting rather than indicating a directional relationship. Nonetheless, the high proportion of respondents reporting delayed nephrology evaluation highlights a potential gap between initial disclosure and structured linkage to specialist care, supporting the need for clearer, standardized post-disclosure pathways.

### Patient organizations and patient-reported evidence

Patient associations played an essential role in the development and dissemination of PK-DIAG, illustrating how community partnerships can generate actionable real-world evidence on diagnostic and care experiences. Large patient-reported registries, such as the ADPKD Registry established by the PKD Foundation, have similarly demonstrated the feasibility and value of capturing patient experience at scale (16,17). Compared with qualitative focus groups involving selected patient experts (18,19)—highly informative for ethical and emotional dimensions but inherently limited in size—PK-DIAG provides complementary insight into real-world diagnostic practices and patient experience across a large sample, allowing evaluation of factors associated with these experiences and downstream associations.

### Strengths and limitations

Strengths include the large patient-centered sample, the close collaboration with patient organizations, and a prespecified de-duplication procedure complemented by manual review. Several limitations warrant consideration. First, recruitment through online patient advocacy channels may introduce selection bias, with overrepresentation of women and individuals with higher educational attainment, potentially limiting generalizability. Second, diagnosis, disclosure circumstances, and emotional responses were self-reported and may be subject to recall bias, particularly for events occurring years earlier. Third, the primary disclosure experience measures—the tact score and the overall diagnostic experience score—were assessed using brief, study-specific single-item patient-reported experience measures that have not undergone full psychometric validation. However, descriptive analyses showed adequate use of the full 0–5 response scale with no marked ceiling effects, supporting their interpretability for capturing extreme negative disclosure experiences in this large sample (Supplementary Table 7). In contrast, health-related quality of life was assessed using the validated ADPKD Impact Scale (ADPKD-IS). Finally, the cross-sectional design precludes causal inference; observed associations may reflect confounding or reverse causality (for example, psychological support around diagnosis may mark greater distress rather than cause poorer experience). Associations between reported post-disclosure intentions and subsequent care timing should therefore be interpreted descriptively. Item-level missingness required complete-case analyses, which may bias estimates if missingness is not random.

### Conclusions

Diagnostic disclosure of ADPKD commonly occurs outside dedicated consultations and is frequently associated with poor tact and unmet psychosocial needs. Very negative diagnostic experiences were strongly linked to modifiable aspects of disclosure quality and information delivery. These findings support implementing structured, guideline-aligned diagnostic disclosure pathways, with systematic counseling and psychosocial support, to better meet patient needs at diagnosis.

## Data Availability

All data produced in the present study are available upon reasonable request to the authors

## Authors contribution

MD and DJ conceived and designed the study. MD, PG, and SSL contributed to questionnaire development and dissemination in collaboration with patient organizations. MD performed data cleaning, deduplication, and statistical analyses, with methodological input from DJ. MD and DJ drafted the initial manuscript. PG, SSL, and SC critically reviewed and revised the manuscript for important intellectual content. All authors approved the final version.

## Acknowledgements

We thank all participants and the patient associations France Polykystose and AIRG-France for their support in developing and disseminating PK-DIAG. We also acknowledge the F-CRIN INI-CRCT network.

## Data Sharing Statement

De-identified participant data will be shared upon reasonable request to the corresponding author, subject to privacy and governance constraints.

## Supplementary material, PKDIAG survey

**Supplementary Figure 1.**
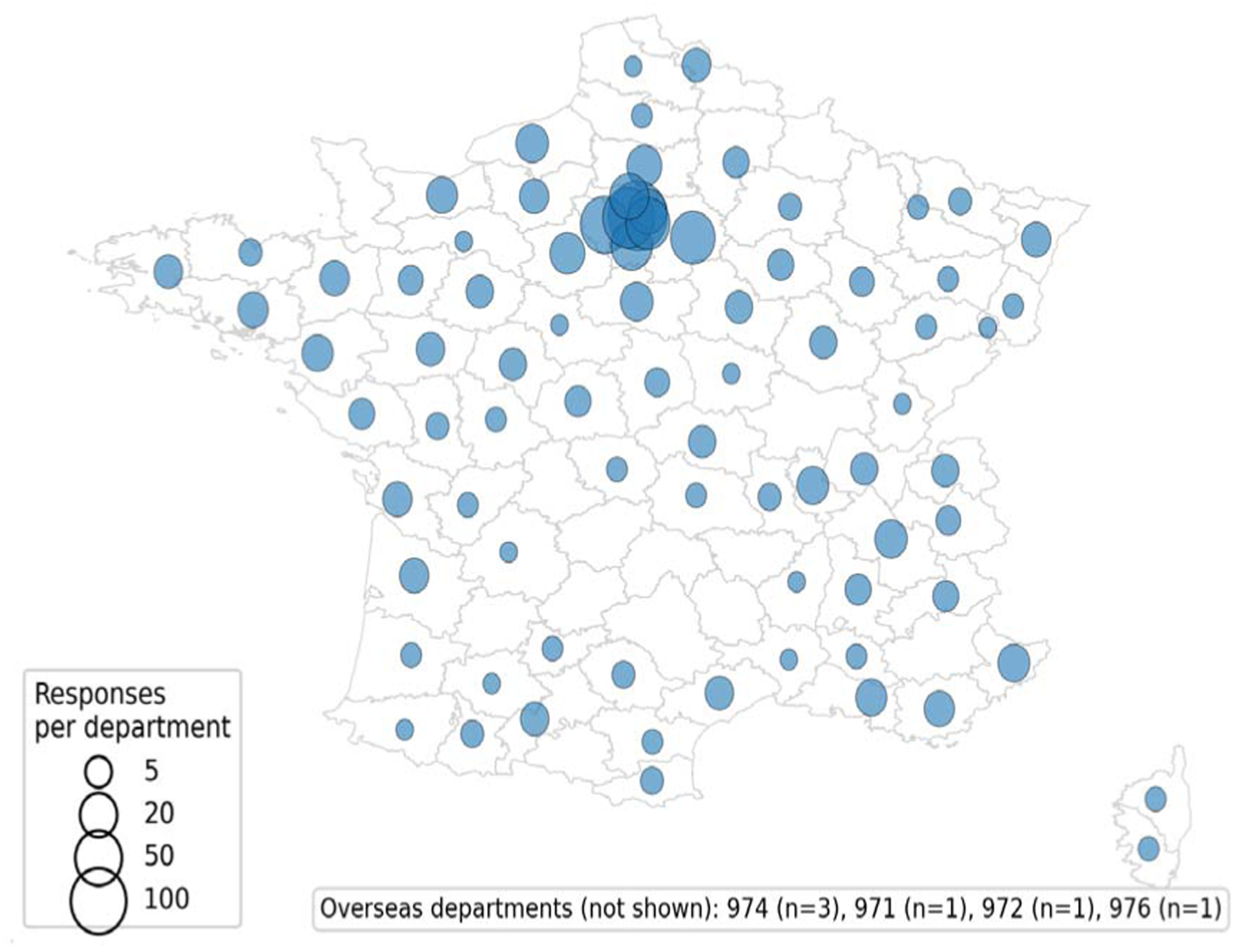
Geographical distribution of PK-DIAG survey respondents. Bubble map showing the self-reported area of residence at the time of the survey, aggregated at the French department level to preserve confidentiality. Circles are centered on each department and scaled proportionally to the number of respondents reporting residence in that department (larger circles indicate higher counts). Department boundaries are shown for reference. Respondents living outside mainland France (overseas territories and/or outside France, depending on reporting) are summarized separately in the Supplementary material. The survey was administered in French; respondents were therefore French speaking.

**Supplementary Table 1.**
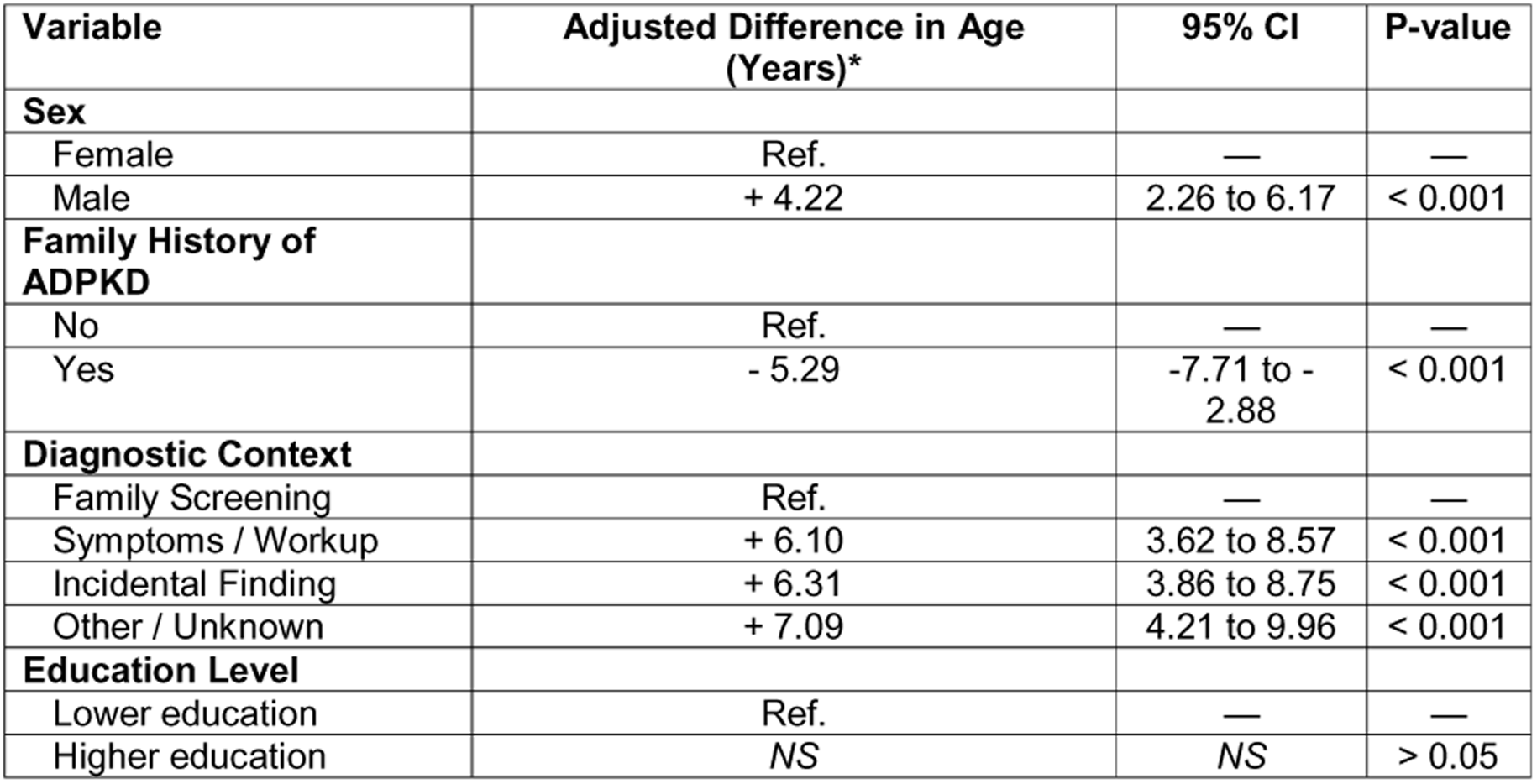
Multivariate linear regression analysis of factors associated with age at ADPKD diagnosis. Coefficients represent the adjusted difference in mean age at diagnosis (in years) compared to the reference group. The model was adjusted for sex, family history, diagnostic context, and education level. Robust standard errors (HC3) were used to account for heteroscedasticity. Complet cases analysis n = 770. CI = Confidence Interval; ADPKD = Autosomal Dominant Polycystic Kidney Disease.

| Variable | Adjusted Difference in Age (Years)* | 95% CI | P-value |
| --- | --- | --- | --- |
| **Sex** |  |  |  |
| Female | Ref. | — | — |
| Male | + 4.22 | 2.26 to 6.17 | < 0.001 |
| **Family History of ADPKD** |  |  |  |
| No | Ref. | — | — |
| Yes | - 5.29 | -7.71 to - 2.88 | < 0.001 |
| **Diagnostic Context** |  |  |  |
| Family Screening | Ref. | — | — |
| Symptoms / Workup | + 6.10 | 3.62 to 8.57 | < 0.001 |
| Incidental Finding | + 6.31 | 3.86 to 8.75 | < 0.001 |
| Other / Unknown | + 7.09 | 4.21 to 9.96 | < 0.001 |
| **Education Level** |  |  |  |
| Lower education | Ref. | — | — |
| Higher education | NS | NS | > 0.05 |

**Supplementary Table 2.** Multivariable logistic regression analysis of factors associated with poor tact diagnostic disclosure. Adjusted odds ratios (aOR) with 95% confidence intervals for poor tact (tact score 0–1 on a 0–5 scale). Model fitted as a complete-case analysis (n=803). Abbreviations: aOR, adjusted odds ratio; CI, confidence interval; ADPKD-IS, ADPKD Impact Scale.

| Variable | aOR | 95% CI |
| --- | --- | --- |
| Age at diagnosis (per 10 years) | 1.00 | 0.87-1.14 |
| Male (vs female) | 0.73 | 0.50-1.05 |
| Higher education (≥Bac+3 vs lower) | 1.50 | 0.92-2.43 |
| **Diagnosis context (reference: family screening)** |  |  |
| Symptoms vs family screening | 1.62 | 1.04-2.52 |
| Incidental vs family screening | 2.11 | 1.36-3.27 |
| Other/unknown vs family screening | 1.58 | 0.91-2.73 |
| **First discloser (reference: nephrologist)** |  |  |
| Radiologist vs nephrologist | 2.50 | 1.57-3.98 |
| Primary care vs nephrologist | 0.44 | 0.19-1.02 |
| Other vs nephrologist | 2.04 | 1.13-3.71 |
| Unknown vs nephrologist | 0.95 | 0.30-3.03 |

**Supplementary Table 3.** Fears expressed at the time of diagnosis (overall and by sex/family history).

| **Fear** | **Overall**<br>**(n=442)** | **Men**<br>**(n=114)** | **Women**<br>**(n=255)** | **FH+**<br>**(n=290)** | **FH-**<br>**(n=85)** |
| --- | --- | --- | --- | --- | --- |
| Dialysis or transplant | 271 (61.3%) | 82 (71.9%) | 154 (60.4%) | 177 (61.0%) | 59 (69.4%) |
| Transmission to children | 210 (47.5%) | 60 (52.6%) | 119 (46.7%) | 143 (49.3%) | 39 (45.9%) |
| Premature death | 85 (19.2%) | 20 (17.5%) | 53 (20.8%) | 56 (19.3%) | 18 (21.2%) |
| Medicalization / frequent visits | 63 (14.3%) | 18 (15.8%) | 36 (14.1%) | 48 (16.6%) | 7 (8.2%) |
| Pain | 49 (11.1%) | 15 (13.2%) | 26 (10.2%) | 29 (10.0%) | 13 (15.3%) |
| None | 59 (13.3%) | 13 (11.4%) | 36 (14.1%) | 40 (13.8%) | 9 (10.6%) |
| Other | 4 (0.9%) | 1 (0.9%) | 3 (1.2%) | 3 (1.0%) | 1 (1.2%) |

**Supplementary Table 4.**
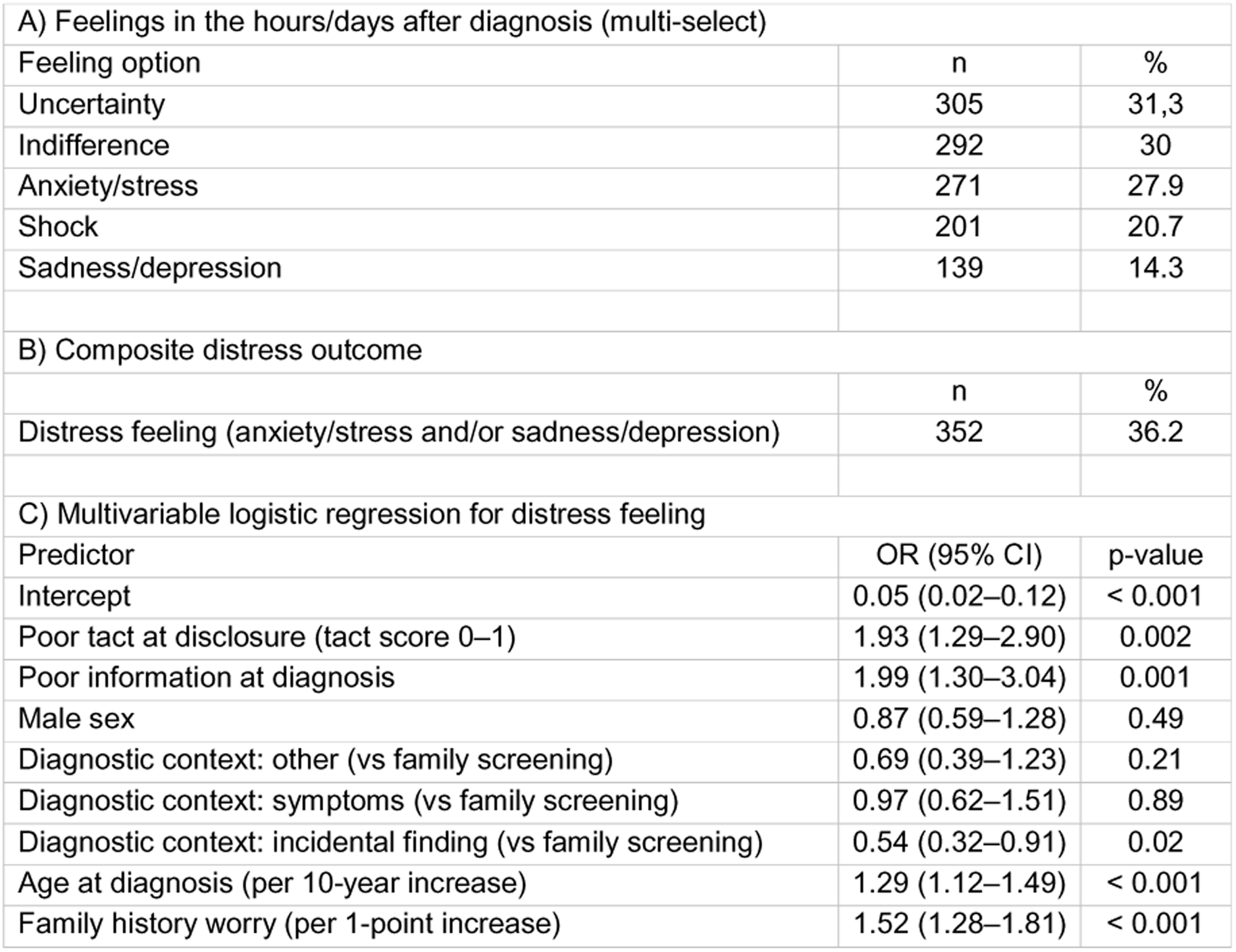
Feelings in the hours/days after diagnosis (multi-select) and factors associated with a composite distress outcome.

**Supplementary Table 5.** Multivariable logistic regression analysis of factors associated with a very negative diagnostic disclosure experience. Complete-case model (N=596). Adjusted odds ratios (aOR) from multivariable logistic regression for very negative experience, defined as a global diagnostic experience score 0–1 on a 0–5 scale (vs 2–5). The model included poor tact, poor information (insufficient/unclear/inaccurate), psychological support at/just after diagnosis (Yes / No but would have been helpful / No not needed [reference]), male sex, higher education, current mood (1–10), ADPKD-IS overall score, and worry about family history (per level). Due to sparse data, the estimate for the “Yes, received” category (n=8) is not displayed in the table or main figure. Abbreviations: aOR, adjusted odds ratio; CI, confidence interval; ADPKD-IS, ADPKD Impact Scale.

| Variable | Adjusted OR (95% CI) | P-value |
| --- | --- | --- |
| **Experience of announcement** |  |  |
| Poor tact / lack of empathy | 4.55 (2.90–7.12) | < 0.001 |
| Poor information quality | 2.52 (1.53–4.15) | < 0.001 |
| **Psychological support** |  |  |
| No, not needed (Reference) | 1.00 (Ref.) | — |
| No, but would have been helpful | 2.60 (1.63–4.17) | < 0.001 |
| **Demographics & context** |  |  |
| Male sex (vs female) | 0.86 (0.53–1.38) | 0.53 |
| Higher education (≥ bachelor) | 0.95 (0.50–1.81) | 0.89 |
| Exploratory adjustment variables (measured at survey) |  |  |
| Mood score (per 1-point) | 0.98 (0.84–1.13) | 0.74 |
| ADPKD-IS overall score (per 1-point) | 0.94 (0.70–1.27) | 0.70 |
| Worry about family history (per level) | 1.37 (1.09–1.71) | 0.01 |

**Supplementary Table 6.** Survey instrument (key items). The survey was administered in French. The table below details the exact wording of the questions and response options used to define the study variables, with their English translation.

| Variable Domain | Question & Response Options (English Translation) | Original Wording (French) |
| --- | --- | --- |
| Diagnostic Context | **In what context was the diagnosis of ADPKD made?**<br>☐ Family screening (asymptomatic)<br>☐ Workup for symptoms (pain, hypertension...)<br>☐ Incidental finding (check-up, surgery)<br>☐ Other<br>☐ Don't know | **Dans quel contexte le diagnostic de polykystose a-t-il été fait ?**<br>☐ Dans le cadre d'un dépistage familial (je n'avais pas de symptôme)<br>☐ Dans le bilan fait à cause de symptômes (douleurs, hypertension...)<br>☐ Découverte fortuite à l'occasion d'un bilan de santé, d'une intervention chirurgicale<br>☐ Autre<br>☐ Je ne sais pas |
| Discloser Specialty | **Who was the first person to tell you that you had polycystic kidney disease?**<br>☐ General Practitioner<br>☐ Nephrologist<br>☐ Radiologist<br>☐ Don't know<br>☐ Other | **Qui a été le premier à vous dire que vous aviez une polykystose ?**<br>☐ Médecin traitant<br>☐ Néphrologue<br>☐ Radiologue<br>☐ Je ne sais pas<br>☐ Autre |
| Disclosure Setting | **In what context did you learn that you had polycystic kidney disease?**<br>☐ Radiology office<br>☐ Medical consultation office<br>☐ Inpatient ward or Emergency Room<br>☐ Hallway or waiting room<br>☐ Self-read report<br>☐ Don't know | **Dans quel contexte avez-vous appris que vous aviez une polykystose ?**<br>☐ Dans le cabinet du radiologue ;<br>☐ En consultation médicale ;<br>☐ En hospitalisation ou aux urgences<br>☐ Dans un couloir ou une salle d'attente<br>☐ En lisant moi-même les résultats sur un compte-rendu<br>☐ Je ne sais pas |
| Tact Score | **In your opinion, how was this diagnostic disclosure made to you?** | **Comment, selon vous, cette annonce diagnostique de polykystose vous a-t-elle été faite ?** |
|  | *(Visual Analog Scale 0–5)*<br>0 = *With no tact at all*<br>5 = *With great tact* | *(Echelle visuelle 0, 1, 2, 3, 4, 5)*<br>0 = *sans aucun tact*<br>5 = *avec un très grand tact* |
| **Global Experience** | **Overall, how did you experience this disclosure?**<br>*(Visual Analog Scale 0–5)*<br>0= extremely bad<br>5= very good | **Globalement, comment avez-vous vécu cette annonce ?**<br>*(Echelle visuelle 0, 1, 2, 3, 4, 5)*<br>0= extrêmement mal<br>5= très bien |
| **Information Quality** | **At the time your diagnosis was made, how do you judge the medical information provided to you?**<br>☐ Impossible to say (did not seek info / too young)<br>☐ Rather clear and understandable information<br>☐ Rather insufficient, incomprehensible or inaccurate information | **Au moment de votre vie où le diagnostic de polykystose a été fait, comment jugez-vous les informations médicales qui vous ont été fournies ?**<br>☐ Impossible à dire (à l'époque je n'ai pas cherché à avoir d'informations /ou j'étais trop jeune)<br>☐ J'ai reçu à l'époque des informations plutôt claires et compréhensibles<br>☐ J'ai reçu à l'époque des informations plutôt insuffisantes, incompréhensibles ou inexacts |
| **Psychological Support** | **At the time of diagnosis or immediately after, did you receive psychological support?**<br>☐ Yes<br>☐ No, I did not need it<br>☐ No, it could have been useful but was not proposed | **Au moment du diagnostic de polykystose, ou dans les suites immédiates, avez-vous bénéficié d'une aide psychologique ?**<br>☐ Oui<br>☐ Non, je n'en ai pas eu besoin<br>☐ Non, cela aurait pu être utile mais on ne me l'a pas proposé |
| **Immediate Feelings** | **What word(s) would best describe your feelings or state of mind in the hours or days following the diagnosis?**<br>☐ Indifference / Carefreeness<br>☐ Shock / Astonishment<br>☐ Sadness / Depression<br>☐ Anxiety / Stress<br>☐ Uncertainty | **Quel(s) mot(s) décrirai(en)t le mieux vos sentiments ou votre état d'esprit dans les heures ou jours qui ont suivi le diagnostic de polykystose ?**<br>☐ Indifférence / Insouciance<br>☐ Choc / étonnement<br>☐ Tristesse / dépression<br>☐ Anxiété / Stress<br>☐ Incertitude |
| **Immediate Resolution** | **After the disclosure of the diagnosis, what resolutions did you take?**<br>☐ Change my lifestyle (drink more, stop smoking...)<br>☐ Initiate specialist follow-up<br>☐ Seek information on the disease by myself<br>☐ None of the above | **Après l'annonce du diagnostic, quelles résolutions avez-vous pris ?**<br>☐ Changer mon mode de vie (boire davantage, ne plus fumer, maigrir...)<br>☐ Entreprendre un suivi spécialisé<br>☐ Me renseigner sur la maladie par moi-même (internet) |
|  |  | ☐ Aucune des résolutions précédentes |
| **Perceived Timing** | **Overall, do you think your diagnosis of polycystic kidney disease was made...**<br>☐ Too late (earlier diagnosis and care would have helped me)<br>☐ At the right time<br>☐ Too early (knowing early was rather negative for me) | **Vous pensez globalement que le diagnostic de votre polykystose a été fait ...**<br>☐ Trop tard (un diagnostic et une prise en charge plus précoces m'auraient rendu service)<br>☐ Au bon moment<br>☐ Trop tôt (jsavoir précocément que j'avais cette maladie a été plutôt négatif pour moi) |
| **Ideal Age** | **At what age do you think it is useful to know if one is affected by polycystic kidney disease in your family?**<br>☐ In childhood<br>☐ In adolescence<br>☐ Between 18 and 30 years old<br>☐ After 30 years old<br>☐ I have no opinion | **A quel âge vous semble-t-il utile de savoir si l'on est atteint de polykystose dans votre famille ?**<br>☐ Dans l'enfance<br>☐ Dans l'adolescence<br>☐ Entre 18 et 30 ans<br>☐ Après 30 ans<br>☐ Je n'ai pas d'avis |

**Supplementary Table 7.** Distributional properties of the tact score and overall diagnostic experience score. The tact score and the overall diagnostic experience score were collected on single-item 0–5 Likert-type scales (0=worst, 5=best). We described their distributions (percentages by response option), median [IQR], and assessed floor/ceiling effects (percentage of responses at the minimum/maximum). In this cohort, both scores used the full response range, with moderate floor effects (tact: 17.2%; experience: 15.6%) and no marked ceiling effects (tact: 13.3%; experience: 11.3%). Tact and overall experience were moderately correlated (Spearman ρ=0.43, p<0.001), consistent with expected convergent validity. Across respondents with both scores available (n=916), the tact score was positively correlated with the overall experience score (Spearman’s ρ = 0.43, p < 0.001). The 0–1 threshold was prespecified to capture the lowest two response categories, reflecting clearly negative disclosure experiences.

|  | **Tact score(n=928)** | **Overall experience score(n=927)** |
| --- | --- | --- |
| **Distribution of scores, n (%)** |  |  |
| Score 0 (worst) | 160 (17%) | 145 (16%) |
| Score 1 | 69 (7%) | 123 (13%) |
| Score 2 | 165 (18%) | 196 (21%) |
| Score 3 | 255 (28%) | 242 (26%) |
| Score 4 | 156 (17%) | 116 (13%) |
| Score 5 (best) | 123 (13%) | 105 (1%) |
| **Summary statistics** |  |  |
| Mean ± SD | 2.6 ± 1.6 | 2.4 ± 1.5 |
| Median [IQR] | 3 [2–4] | 2 [1–3] |
| Distribution characteristics |  |  |
| Floor effect (score=0) | 17% | 16% |
| Ceiling effect (score=5) | 13% | 11% |
| Worst two categories (scores 0–1) | 25% | 29% |
| Best two categories (scores 4–5) | 30% | 24% |

## REFERENCES

1. Chebib FT, Hanna C, Harris PC, Torres VE, Dahl NK: Autosomal Dominant Polycystic Kidney Disease: A Review. JAMA 333: 1708–1719, 2025

2. Tong A, Rangan GK, Ruospo M, Saglimbene V, Strippoli GFM, Palmer SC, Tunnicliffe DJ, Craig JC: A painful inheritance-patient perspectives on living with polycystic kidney disease: thematic synthesis of qualitative research. Nephrol Dial Transplant 30: 790–800, 2015

3. Ebrahimi N, Garimella PS, Chebib FT, Sparks MA, Lerma EV, Golsorkhi M, Ghozloujeh ZG, Abdipour A, Norouzi S: Mental Health and Autosomal Dominant Polycystic Kidney Disease: A Narrative Review. Kidney360 [Internet] 5: 1200, 2024 Available from: https://journals.lww.com/kidney360/fulltext/2024/08000/mental_health_and_autosomal_dominant_polycystic.21.aspx [cited 2026 Jan 22]

4. Mustafa et al. RA, Kawtharany, H, Kalot, MA: Meaningful patient-centered outcomes in PKD: priorities for future research. 2025 Available from: https://journals.lww.com/kidney360/fulltext/2025/04000/establishing_meaningful_patient_centered_outcomes.15.aspx

5. Logeman C, Cho Y, Sautenet B, Rangan GK, Gutman T, Craig J, Ong A, Chapman A, Ahn C, Coolican H, Tze-Wah Kao J, Gansevoort RT, Perrone R, Harris T, Torres V, Fowler K, Pei Y, Kerr P, Ryan J, Johnson D, Viecelli A, Geneste C, Kim H, Kim Y, Howell M, Ju A, Manera KE, Teixeira-Pinto A, Parasivam G, Tong A: “A sword of Damocles”: patient and caregiver beliefs, attitudes and perspectives on presymptomatic testing for autosomal dominant polycystic kidney disease: a focus group study. BMJ Open 10: e038005, 2020

6. Kidney Disease: Improving Global Outcomes (KDIGO) ADPKD Work Group: KDIGO 2025 Clinical Practice Guideline for the Evaluation, Management, and Treatment of Autosomal Dominant Polycystic Kidney Disease (ADPKD). Kidney Int 107: S1–S239, 2025

7. Forbes Shepherd R, Browne TK, Warwick L: A Relational Approach to Genetic Counseling for Hereditary Breast and Ovarian Cancer. J Genet Couns 26: 283–299, 2017

8. Gimpel C, Fieuws S, Hofstetter J, Pitcher D, Vanmeerbeek L, Haeberle S, Dachy A, Massella L, Seeman T, Ranchin B, Allard L, Bacchetta J, Bayrakci US, Becherucci F, Perez-Beltran V, Besouw M, Bialkevich H, Boyer O, Canpolat N, Chauveau D, Çiçek N, Conlon PJ, Devuyst O, Dossier C, Fila M, Flögelová H, Godron-Dubrasquet A, Gokce I, Nguyen-Tang EG, González-Rodríguez JD, Guffens A, Grandaliano G, Heidet L, Jankauskiene A, Levart TK, Knebelmann B, König JC, La Scola C, Leone VF, Leroy V, Litwin M, Lucchetti L, Lungu AC, Marzuillo P, Mastrangelo A, Miklaszewska M, Montini G, Nobili F, Obrycki L, Papizh S, Paripović A, Paripović D, Peruzzi L, Raes A, Saygili S, Spasojević B, Simon T, Szczepańska M, Trepiccione F, Varda NM, Westland R, Yüksel S, Zaluska-Lesniewska I, Tenebaum J, Mustafa R, Mallett AJ, Guay-Woodford LM, Gale DP, Böckenhauer D, Liebau MC, Schaefer F, Mekahli D, RaDaR ADPKD Rare Disease Group, ERKReg Collaborators, ADPedKD Collaborators: Insights from ADPedKD, ERKReg and RaDaR registries provide a multi-national perspective on the presentation of childhood autosomal dominant polycystic kidney disease in high- and middle-income countries. Kidney Int 108: 105–118, 2025

9. Baker A, King D, Marsh J, Makin A, Carr A, Davis C, Kirby C: Understanding the physical and emotional impact of early-stage ADPKD: experiences and perspectives of patients and physicians. Clin Kidney J 8: 531–537, 2015

10. Gimpel C, Bergmann C, Bockenhauer D, Breysem L, Cadnapaphornchai MA, Cetiner M, Dudley J, Emma F, Konrad M, Harris T, Harris PC, König J, Liebau MC, Marlais M, Mekahli D, Metcalfe AM, Oh J, Perrone RD, Sinha MD, Titieni A, Torra R, Weber S, Winyard PJD, Schaefer F: International consensus statement on the diagnosis and management of autosomal dominant polycystic kidney disease in children and young people. Nat Rev Nephrol 15: 713–726, 2019

11. Simms RJ, Thong KM, Dworschak GC, Ong ACM: Increased psychosocial risk, depression and reduced quality of life living with autosomal dominant polycystic kidney disease. Nephrol Dial Transplant [Internet] 31: 1130–1140, 2016 Available from: 10.1093/ndt/gfv299 [cited 2026 Jan 10]

12. Cho Y, Sautenet B, Gutman T, Rangan G, Craig JC, Ong AC, Chapman A, Ahn C, Coolican H, Kao JT-W, Gansevoort R, Perrone RD, Harris T, Torres V, Pei Y, Kerr PG, Ryan J, Johnson DW, Viecelli AK, Geneste C, Kim H, Kim Y, Oh YK, Teixeira-Pinto A, Logeman C, Howell M, Ju A, Manera KE, Tong A: Identifying patient-important outcomes in polycystic kidney disease: An international nominal group technique study. Nephrology (Carlton) 24: 1214–1224, 2019

13. Oberdhan MJ D, Cole, JC,& Atkinson: Development of a patient-reported outcomes tool to assess pain and discomfort in autosomal dominant polycystic kidney disease. 2023 Available from: https://journals.lww.com/cjasn/fulltext/2023/02000/development_of_a_patient_reported_outcomes_tool_to.12.aspx

14. Natale P, Hannan E, Sautenet B, Ju A, Perrone RD, Burnette E, Casteleijn N, Chapman A, Eastty S, Gansevoort R, Hogan M, Horie S, Knebelmann B, Lee R, Mustafa RA, Sandford R, Baumgart A, Tong A, Strippoli GFM, Craig JC, Rangan GK, Cho Y: Patient-reported outcome measures for pain in autosomal dominant polycystic kidney disease: A systematic review. PLoS One [Internet] 16: e0252479, 2021 Available from: https://www.ncbi.nlm.nih.gov/pmc/articles/PMC8158964/ [cited 2022 Feb 11]

15. Joly K D, Quinn, J, Mokiou, S,& O’Reilly: Patient experience and satisfaction in ADPKD: a European cross-sectional study. 2020 Available from: 10.1186/s12882-020-01927-1

16. Hoover B E, Perrone, RD, Rusconi, C,& Benson: National patient-powered registry in ADPKD: design & implementation. 2022 Available from: https://journals.lww.com/kidney360/fulltext/2022/08000/design_and_basic_characteristics_of_a_national.12.aspx

17. Hoover D E, Holliday, V, Merullo, N,& Oberdhan: Pain and health-related quality of life in ADPKD: results from a national patient-powered registry. 2024 Available from https://www.sciencedirect.com/science/article/pii/S2590059524000244

18. Harris T, Bridges HR, Brown WD, O’Brien NL, Daly AC, Jindal BK, Mundy GS, Ong A, Power AJ, Sandford RN, Sayer J, Simms RJ, Wilson PD, Winyard PJD, Tarpey M: Research priorities for autosomal dominant polycystic kidney disease: a UK priority setting partnership. BMJ Open 12: e055780, 2022

19. Gosselink ME, Mooren R, Snoek R, Crombag NMTH, Vos P, Keijzer-Veen MG, van Eerde AM, Lely AT: Perspectives of Patients and Clinicians on Reproductive Health Care and ADPKD. Kidney Int Rep 9: 3190–3203, 2024

20. Andreeva VA, Allès B, Feron G, Gonzalez R, Sulmont-Rossé C, Galan P, Hercberg S, Méjean C: Sex-Specific Sociodemographic Correlates of Dietary Patterns in a Large Sample of French Elderly Individuals. Nutrients 8: 484, 2016

